# Validation of real-world actigraphy to capture post-stroke motor recovery

**DOI:** 10.1101/2024.11.03.24316674

**Authors:** Keith R. Lohse, Allison E. Miller, Marghuretta D. Bland, Jin-Moo Lee, Catherine E. Lang

**Affiliations:** Program in Physical Therapy, Washington University School of Medicine; Department of Neurology, Washington University School of Medicine; Division of Brain and Biological Sciences, Washington University School of Medicine; Program in Occupational Therapy, Washington University School of Medicine

## Abstract

Stroke is a leading cause of long-term disability, but advances for rehabilitation have lagged those for acute treatment. Large biological studies (e.g., “omics”-based approaches) may offer mechanistic insights for recovery, but to enable those studies, researchers need to collect detailed recovery phenotypes at scale, e.g., in thousands of people with minimal burden for participants and researchers. This study investigates the concurrent validity between remotely collected wearable sensor data and clinical assessments of motor recovery post-stroke. We specifically focus on the “use ratio”, which is the activity level of the paretic arm relative to the non-paretic arm, measured via bilateral wrist-worn accelerometers. Utilizing a large, harmonized multi-site dataset of adults with stoke, we analyzed cross-sectional (N=198) and longitudinal (N=98) changes in use ratio, the Action Research Arm Test (ARAT) and the Fugl-Meyer Assessment upper extremity subscale (FM-UE). Our findings indicate strong concurrent validity of the use ratio and the ARAT, and the use ratio and the FM-UE both cross-sectionally (i.e., *differences* between people) and longitudinally (i.e., *changes* within a person). Notably, while the use ratio strongly correlated with FM-UE and ARAT initially, the strength of these correlations reduced over time. This decreasing correlation might be explained by the increasing influence that personal and environmental factors play as recovery progresses. Additionally, these correlations were also stronger for the use ratio than for hours of activity for the paretic/nonparetic arm alone, suggesting that it is specifically asymmetry of activity that correlates with clinical measures. Thus, the use ratio is an efficient and clinically valid measure of motor recovery post-stroke that can be deployed at scale to collect biologically meaningful phenotypes.

## Introduction

Stroke is a leading cause of long-term disability worldwide.^1^ While major advances have been made for the treatment of acute ischemic stroke, innovations for recovery and rehabilitation following stroke remain more limited.^2^ Notably, recent clinical trials in stroke rehabilitation yielded neutral results^3–5^ and longitudinal studies show tremendous heterogeneity in both patients’ trajectories and endpoints for recovery.^6,7^ To improve the efficacy of stroke rehabilitation, we need a better understanding of the biological mechanisms underlying recovery, either to develop new interventions or to better match existing interventions to the most responsive patients.^8,9^ “Omics”-based approaches offer a fruitful avenue for gaining this understanding but require data collection on a scale generally not seen in stroke rehabilitation research; e.g, “large” trials in stroke rehabilitation collect data from N<400 people^4^, compared to the (tens of) thousands needed for genome-wide association studies.^10,11^

It is not sufficient to collect large numbers of biospecimens, however, if we do not also have good behavioral phenotypes to define recovery.^12^ Detailed clinical phenotypes are thus generally preferable to proxy measures because they are more likely to capture biological mechanisms. However, these detailed assessments are also costly and difficult to deploy at scale. For instance, the Fugl-Meyer Assessment^13^ of motor recovery (a common tool used in motor recovery studies) takes about 30 minutes to administer. This time *multiplied by* the number of patients *multiplied by* the number of assessments per patient, plus travel time for the patient into the clinic, make these kinds of assessment costly and time consuming. Other clinical measures, like the modified Rankin Scale, are arguably more scalable, but are cruder and can have undesirable measurement properties, such as floor/ceiling effects.^14^ Similarly, patient self-report measures are very valuable and scalable, but fundamentally further from biology, as self-report is inherently filtered through a patient’s own perceptions and often tracks more closely with measures of participation than with underlying impairments in body structure/function.^15,16^ Thus, there is a need for a scalable metric that can capture biologically meaningful phenotypes.

To that end, the goal of the present study was to understand how scalable wearable sensor data correlates with less scalable, but well-validated clinical assessments, and how these correlations change over time after stroke. Specifically, we focused on the *use ratio* for the paretic arm relative to the non-paretic arm collected through bilateral, wrist worn accelerometers. The use ratio is an excellent candidate measure given that: (1) it has an analogue in basic research where fore-paw asymmetry is used as an outcome in rodent models of stroke recovery^17,18^; (2) human research shows the use ratio is feasibly collected in adults at all stages post-stroke^19^, (3) in neurologically intact human adults, the use ratio is narrowly distributed around 1.0 but does not have a hard ceiling like many in-clinic assessments^20^; and (4) the use ratio is an objective “real-world” measure collected passively during daily life, reducing the burden on patients and clinicians.^21–23^ Our in-clinic measures were the upper extremity subscale of the Fugl-Meyer Assessment, which is considered a measure of *body function/structure* within the International Classification of Functioning, Disability, and Health (ICF) framework^24,25^, and the Action Research Arm Test, which is a measure of *activity capacity* within the ICF framework.^24,26^ These data offer new insights on when and how scalable technologies, specifically wearable sensors, can be deployed to measure motor recovery in individuals who have had a stroke.

## Methods

### Data Sources

Measurement data for this study were taken from a large, harmonized dataset of wearable sensor data^27^ hosted as two studies on the National Center for Child and Human Development (NICHD) Data and Specimen Hub (DASH) repository.^21,22^ The overall dataset contains 2,885 days of recording from 790 individuals from numerous populations (e.g., 46% neurotypical, 31% adults with stroke, 7% adults with Parkinson disease), collected from several study sites. Our analyses focused only on adults with stroke who had *at least* some accelerometry data with concurrent data from clinical assessments. Specifically, we analyzed the data in two subsets: a cross-sectional subset (N=198) and a longitudinal subset (N=98).

For the *cross-sectional subset*, we considered participants who were enrolled at any time following their stroke but used only one observation per person in any time window, taking the earliest available observation in [0, 6), [6, 12), [12, 24), [24, 36), [36, 52) and ≥52 weeks; see Figure 1A. (Note ‘[‘ indicates the value is included whereas ‘)’ indicates the point is excluded from that set.) For the *longitudinal subset*, we only considered participants who were enrolled <12 weeks after stroke and modeled the trajectory of their recovery across all available time points, see trajectories for example participants in Figure 1B and 1C.

**Figure 1.**
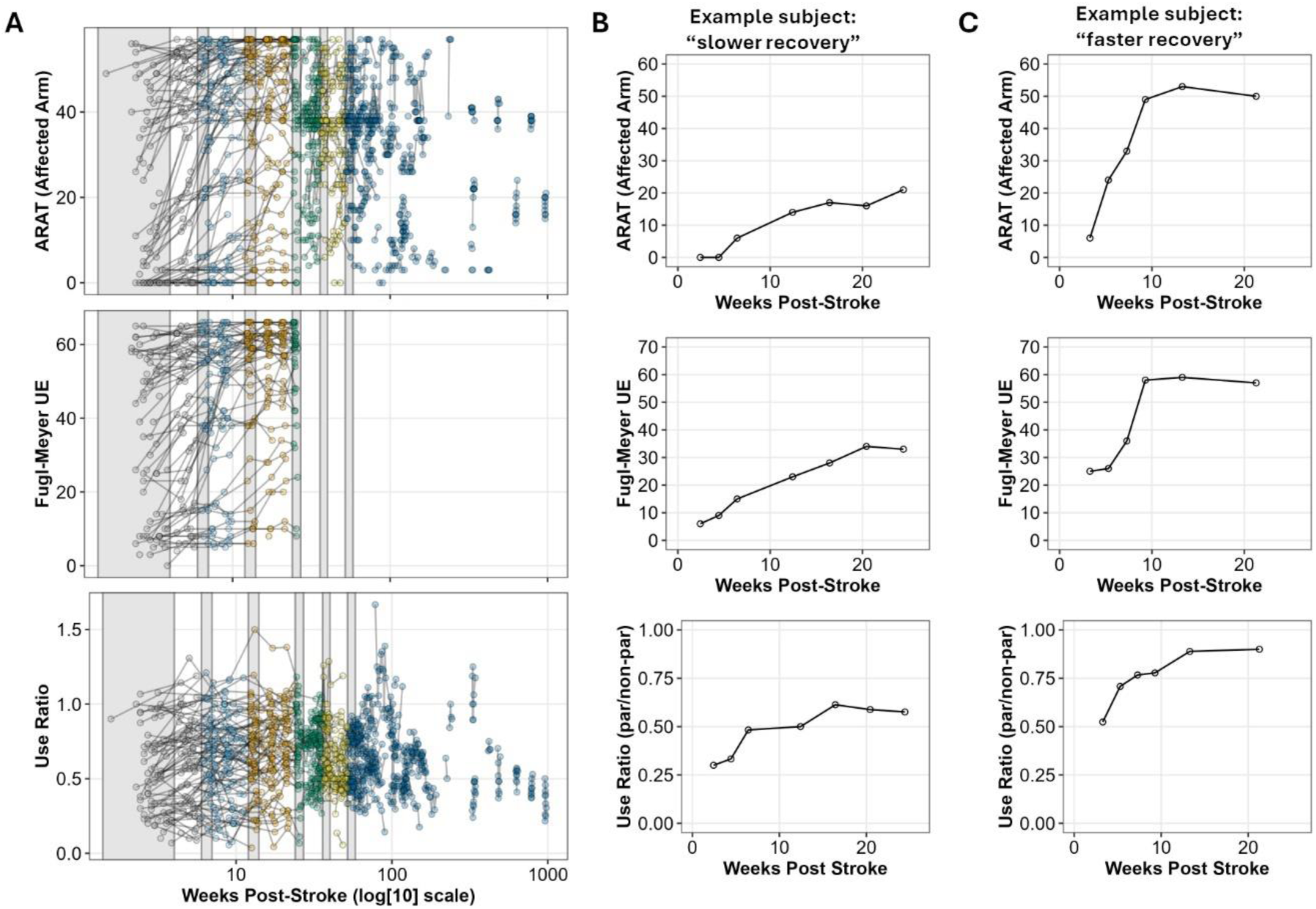
(**A**) An overview of when data were available in our pooled observational subset. Points are color coded by *a priori* bins of time in weeks: [0, 6); [6, 12); [12, 24); [24, 36); [36, 52) and ≥52 weeks. For cross-sectional analyses, we took the first observation available for a participant in each bin, approximated by the grey-shaded areas. (**B and C**) Example data from a participant who showed slower and less recovery (**B**), and a participant who showed faster and more pronounced recovery (**C**) across our three primary outcomes: the ARAT, the FM-UE, and the Use Ratio.

### Primary Outcomes

#### In-Clinic Outcomes

The Fugl-Meyer assessment is the most widely used tool to quantify impairments in motor control after stroke. The upper extremity subscale (FM-UE) includes an assessment of the individual’s reflexes and ability to execute various movement patterns, where higher scores are better (range 0-66).^13,28^ Because the FM-UE assesses fundamental sensorimotor abilities (e.g., volitional movements with and without synergies, coordination, reflexes), it is considered to be a measure of *body function* within the ICF framework.^24,29^ The Action Research Arm Test (ARAT) is a widely used measure of post-stroke upper limb *activity capacity* (i.e., what a person can do on a standardized assessment in the clinic). Clinicians rate the individual’s ability to complete 19 tasks, such as reaching, grasping, and manipulating everyday objects, where higher scores are better (range 0-57).^29,30^ Because the items of the ARAT require the combination of different sensorimotor functions in order to successfully complete various tasks within a structured environment, the ARAT is considered to be a measure of *activity capacity* in the ICF framework.^24,28^

#### Remote Outcomes

Bilateral wrist accelerometers were worn for 1-3 days per time-point, as defined in the original study protocols.^19,31–33^ The wrist accelerometers were either the ActiGraph GT3X-BT or GT9X-Link device (ActiGraph, Pensacola, FL). These devices includes a tri-axial accelerometer and sampled at 30 Hz. To compute the use ratio, data were first filtered in ActiLife 6 software using its proprietary algorithm, which uses a maximum gain of 0.759 Hz and goes down to −6db at 0.212 Hz and 2.148 Hz.^34^ Data were then converted to activity counts by summing the down-sampled signal within 1-second epochs for each axis. A 1-Hz data file (in activity counts) was extracted from ActiLife 6 software and imported into R (R Core Team, 2013, version 4.2.1) for further processing^35^ using custom-written code (available at: https://github.com/keithlohse/HarmonizedAccelData^36^). Activity counts in each axis were combined into a single vector magnitude using the formula 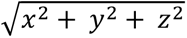. A threshold of ≥ 2 activity counts was used to determine if the upper limb was active for each second.^37,38^ The number of hours of paretic upper limb activity was calculated by summing the seconds of movement of the paretic upper limb that exceeded the threshold and converting to hours. The same process was done to compute the number of hours of non-paretic upper limb activity. These two variables were then used to compute the use ratio as the hours of paretic limb movement relative to the hours of non-paretic limb movement.^39^ The use ratio is considered a measure of *activity performance* under the ICF framework, because it captures activity of the upper extremities in daily life, outside of a structured clinical environment.^19,24,30^

When interpreting the use ratio as an outcome, it might be regarded as a relatively simple metric. Especially given the wide range of possible variables that can be extracted from the accelerometer data. However, there is an elegance and robustness to this simplicity. For instance, by using a low threshold to classify periods of “movement” versus “no movement”, the use ratio does not rely on the classification of “functional” versus “non-functional” movements, which is an area of disagreement in the field. For example, some authors have considered the hold time during grasping as non-functional, along with any upper limb movements when a person is walking/ or moving around in a space.^40^ However, during activities of daily living, people hold objects with one hand while manipulating the object with another, either while stationary or while moving (e.g. meal preparation in the kitchen). The threshold used here includes all upper limb movement in the numerator and denominator, agnostic to the classification of functional vs. non-functional. Similarly, the calculation of the use ratio in people with stroke appears to be robust to a number of analytical choices in the processing pipeline. For instance, use ratio variables are minimally influenced by including or excluding sleep in the data processing pipeline (sd of difference = 0.018) in non-disabled adults and adults with upper limb disability, including those with stroke measured during daily life.^41^

### Statistical Analysis

#### Cross-Sectional Associations

Within each time window, we took only the first available observation for each participant in that window (creating independent data within each window). In each window, we then calculated the Pearson correlation between the paretic and non-paretic limbs, between the use ratio and the ARAT, and between the use ratio and the FM UE. This led to the following number of complete pairs at different times for the use ratio and the ARAT: [0, 6) n=81; [6, 12) n=85; [12, 24) n=72; [24, 36) n=68; [36, 52) n=38 and ≥52 n=66. And for the use ratio and the FM UE: [0, 6) n=67; [6, 12) n=57; [12, 24) n=49; [24, 36) n=37; [36, 52) n=0 and ≥52 n=0.

#### Longitudinal Associations

Based on previous work^7,19,42^ and visual inspection for the trajectories of different participants, we saw strong evidence of nonlinear trajectories for the ARAT, FM-UE, and the use ratio. To capture this change, we tested a series of curvilinear and nonlinear mixed-effect models^43,44^ with Time (in weeks post stroke) included as a fixed effect. Random effects included random intercepts for each participant and random slopes for the different versions of the time variable (depending on the model). Details of the model fits are shown in Supplemental Tables i-iii, but in brief, across all outcomes, the best fitting model was a 2-knot linear spline. The placement of the knots was determined by comparing different knot locations in a grid search and comparing models based on Akaike’s Information Criterion (AIC). As summarized in Supplemental Figure i, the best fitting knots were generally around 4-7 weeks and 10-13 weeks for the ARAT, FM-UE, and Use Ratio. The minimum AIC was not in the exact same location for all outcomes, but knots at weeks 5 and 11 yielded some of the smallest AICs across outcomes and correspond to clinically meaningful time points in recovery.^45,46^ Thus, we chose to fit splines with knots at 5 and 11 weeks for all models, see Figure 2.

**Figure 2.**
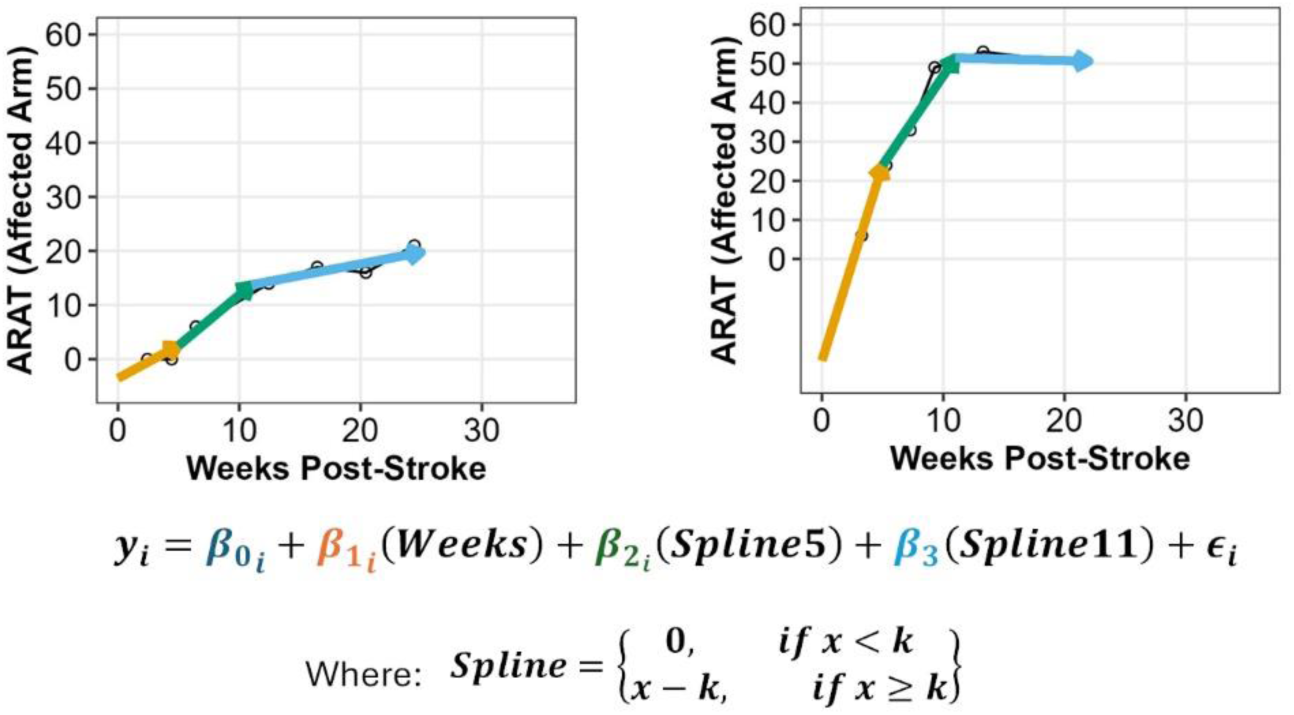
An illustration of the flexibility of a 2-knot spline model. The same model is fit to data from two different participants. Note that the intercept reflects the hypothetical value of the outcome when a person is at 0 weeks post-stroke. As seen in both participants, this intercept can be negative even when the outcome has a floor at 0, so intercepts should be interpreted with caution. x = time in weeks, k=location of the knot in weeks.

We note that 5 and 11 weeks are clearly not the absolute nor the only clinically meaningful inflection points in recovery, but these two points do capture clinically meaningful change in the three outcomes analyzed here and are reasonably close to the 1 and 3 month time points seen in older epidemiological recovery studies.^47,48^ By putting the knots in the same location, we can make an “apples to apples” comparison of trajectories across models. Specifically, by extracting the unique coefficients for each person as shown in Figure 2, we can calculate *an intercept* that represents their estimated value at Week 0. And, by taking the partial derivative with respect to time, we can solve for each person’s unique slope before and after the two different knots: i.e., at Week 0 = *β*_1_; at Week 5 = (*β*_1_ + *β*_2_); and at Week 11 = (*β*_1_ + *β*_2_ + *β*_3_). These slopes reflect the *change per week* for a given participant in that time window. We then calculated Pearson correlations and 95% confidence intervals for regression terms between the use ratio and ARAT and the use ratio and FM-UE. For completeness, we also compared the trajectories for the ARAT compared to the FM-UE. These correlations provide context for the multivariate nature of recovery, but do not directly address the validity of the use ratio.

Unadjusted 95% confidence intervals are presented for all correlations, with *r*^2^ values presented in the figures. Note that our focus is not on the statistical significance of these correlations per se, but rather the magnitude and precision of the estimate (e.g., r=0.25 is statistically significant at our sample size, but suggests limited concurrent validity between measures, as the *r*^2^ would only be 0.06 or 6% of the variance explained). Code for all data processing and analyses is available from: https://github.com/keithlohse/useRatio_validityStudy, and data are available from NICHD DASH.^21,22^

## Results

### 1. Cross Sectional Associations between In-Clinic Variables and the Use Ratio

Descriptive statistics for the cross-sectional subset are provided in Table 1. Broadly, this sample consisted of a largely Black and White, non-Hispanic, older adult cohort who had experienced their first stroke, predominately cortical ischemic strokes. At enrollment, 50% of participants were <19 weeks post-stroke. Participants also tended to have mild to moderate cognitive impairment, and a wide range of motor impairment for the upper extremities, ranging from mild to very severe.

**Table 1.**
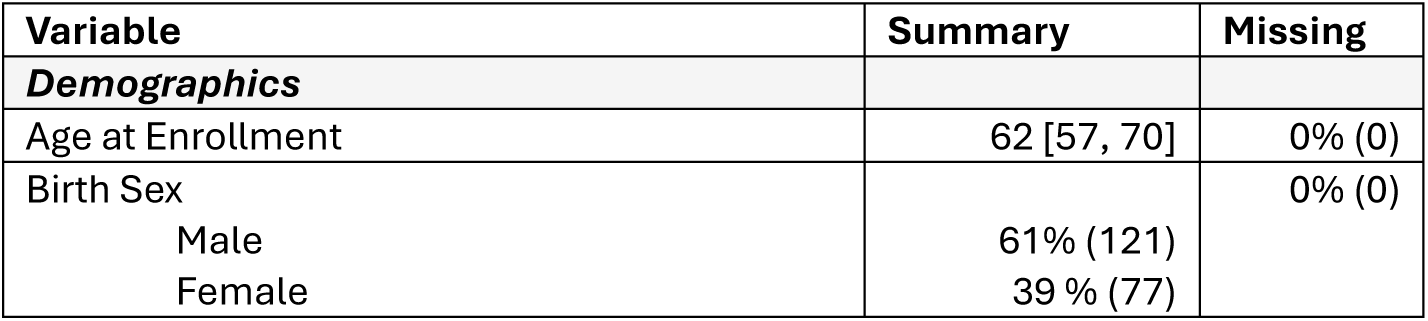
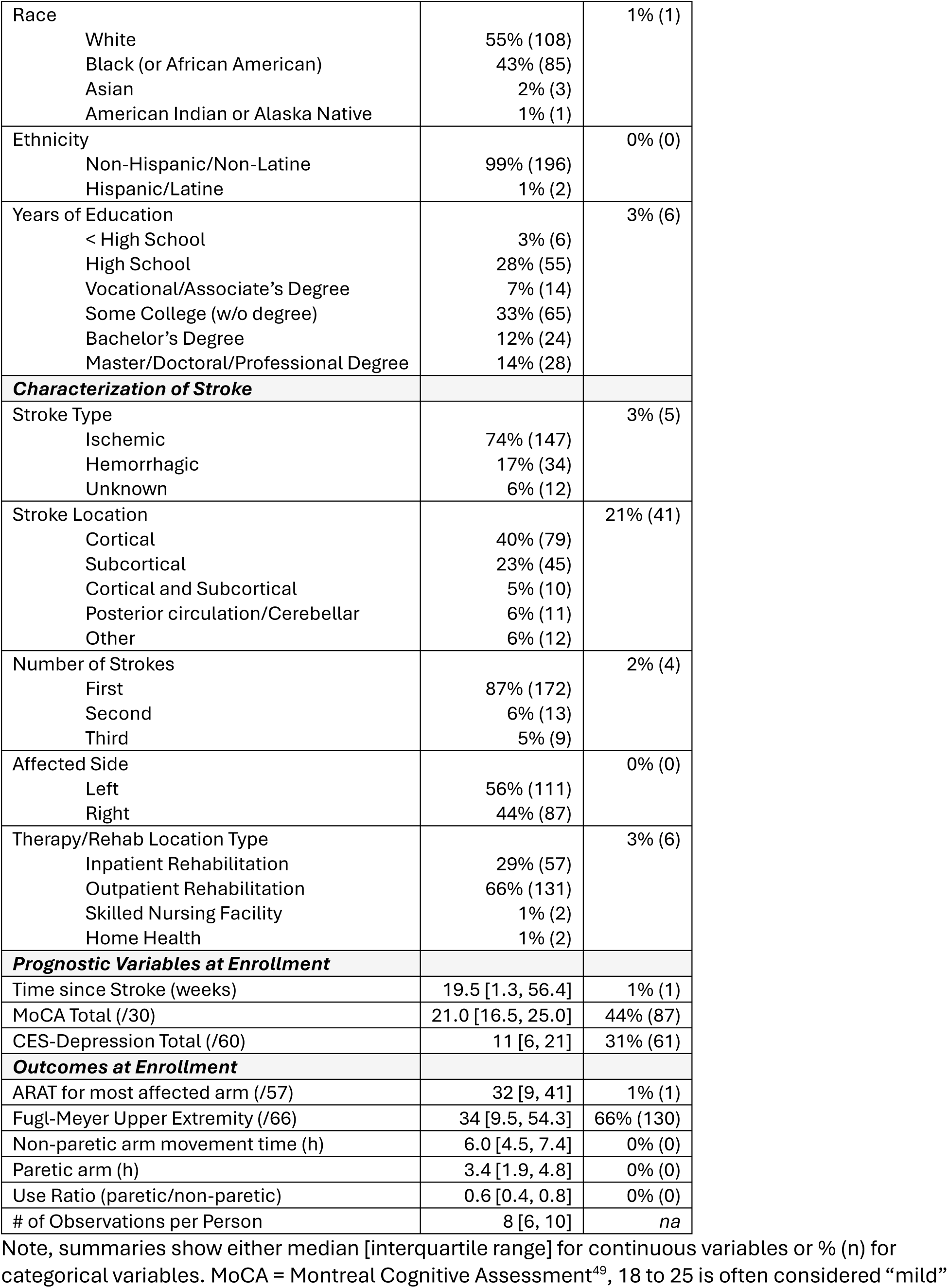
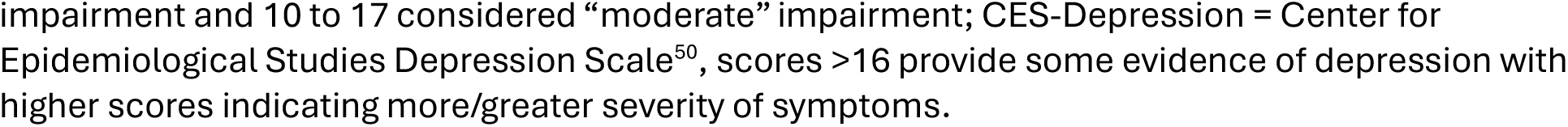
Descriptive statistics for the cross-sectional cohort (N=198).

| Variable | Summary | Missing |
| --- | --- | --- |
| **Demographics** |  |  |
| Age at Enrollment | 62 [57, 70] | 0% (0) |
| Birth Sex |  | 0% (0) |
| Male | 61% (121) |  |
| Female | 39 % (77) |  |
| Race |  | 1% (1) |
| White | 55% (108) |  |
| Black (or African American) | 43% (85) |  |
| Asian | 2% (3) |  |
| American Indian or Alaska Native | 1% (1) |  |
| Ethnicity |  | 0% (0) |
| Non-Hispanic/Non-Latine | 99% (196) |  |
| Hispanic/Latine | 1% (2) |  |
| Years of Education |  | 3% (6) |
| < High School | 3% (6) |  |
| High School | 28% (55) |  |
| Vocational/Associate's Degree | 7% (14) |  |
| Some College (w/o degree) | 33% (65) |  |
| Bachelor's Degree | 12% (24) |  |
| Master/Doctoral/Professional Degree | 14% (28) |  |
| **Characterization of Stroke** |  |  |
| Stroke Type |  | 3% (5) |
| Ischemic | 74% (147) |  |
| Hemorrhagic | 17% (34) |  |
| Unknown | 6% (12) |  |
| Stroke Location |  | 21% (41) |
| Cortical | 40% (79) |  |
| Subcortical | 23% (45) |  |
| Cortical and Subcortical | 5% (10) |  |
| Posterior circulation/Cerebellar | 6% (11) |  |
| Other | 6% (12) |  |
| Number of Strokes |  | 2% (4) |
| First | 87% (172) |  |
| Second | 6% (13) |  |
| Third | 5% (9) |  |
| Affected Side |  | 0% (0) |
| Left | 56% (111) |  |
| Right | 44% (87) |  |
| Therapy/Rehab Location Type |  | 3% (6) |
| Inpatient Rehabilitation | 29% (57) |  |
| Outpatient Rehabilitation | 66% (131) |  |
| Skilled Nursing Facility | 1% (2) |  |
| Home Health | 1% (2) |  |
| **Prognostic Variables at Enrollment** |  |  |
| Time since Stroke (weeks) | 19.5 [1.3, 56.4] | 1% (1) |
| MoCA Total (/30) | 21.0 [16.5, 25.0] | 44% (87) |
| CES-Depression Total (/60) | 11 [6, 21] | 31% (61) |
| **Outcomes at Enrollment** |  |  |
| ARAT for most affected arm (/57) | 32 [9, 41] | 1% (1) |
| Fugl-Meyer Upper Extremity (/66) | 34 [9.5, 54.3] | 66% (130) |
| Non-paretic arm movement time (h) | 6.0 [4.5, 7.4] | 0% (0) |
| Paretic arm (h) | 3.4 [1.9, 4.8] | 0% (0) |
| Use Ratio (paretic/non-paretic) | 0.6 [0.4, 0.8] | 0% (0) |
| # of Observations per Person | 8 [6, 10] | na |

Note, summaries show either median [interquartile range] for continuous variables or % (n) for categorical variables. MoCA = Montreal Cognitive Assessment<sup>49</sup>, 18 to 25 is often considered “mild”
impairment and 10 to 17 considered “moderate” impairment; CES-Depression = Center for Epidemiological Studies Depression Scale<sup>50</sup>, scores >16 provide some evidence of depression with higher scores indicating more/greater severity of symptoms.

First, there were generally positive correlations between the hours of activity for the paretic limb and the non-paretic limb (Figure 3A). As expected, time for the paretic limb was generally lower than the non-paretic limb, but there was a similar correlation between limbs across the various time points: at [0, 6) weeks r = 0.72 and 95% CI=[0.59, 0.81]; at [6, 12) weeks r = 0.79 and 95% CI=[0.69, 0.86]; at [12, 24) weeks r = 0.77 and 95% CI=[0.66, 0.85]; at [24, 36) weeks r= 0.81 and 95% CI=[0.71, 0.88]; at [36, 52) weeks r = 0.80 and 95% CI=[0.64, 0.89]; and at ≥52 weeks r= 0.70 and 95% CI=[0.55, 0.81]. Thus, the use ratio captures a similar relationship between the paretic and non-paretic limb activity over time.

**Figure 3.**
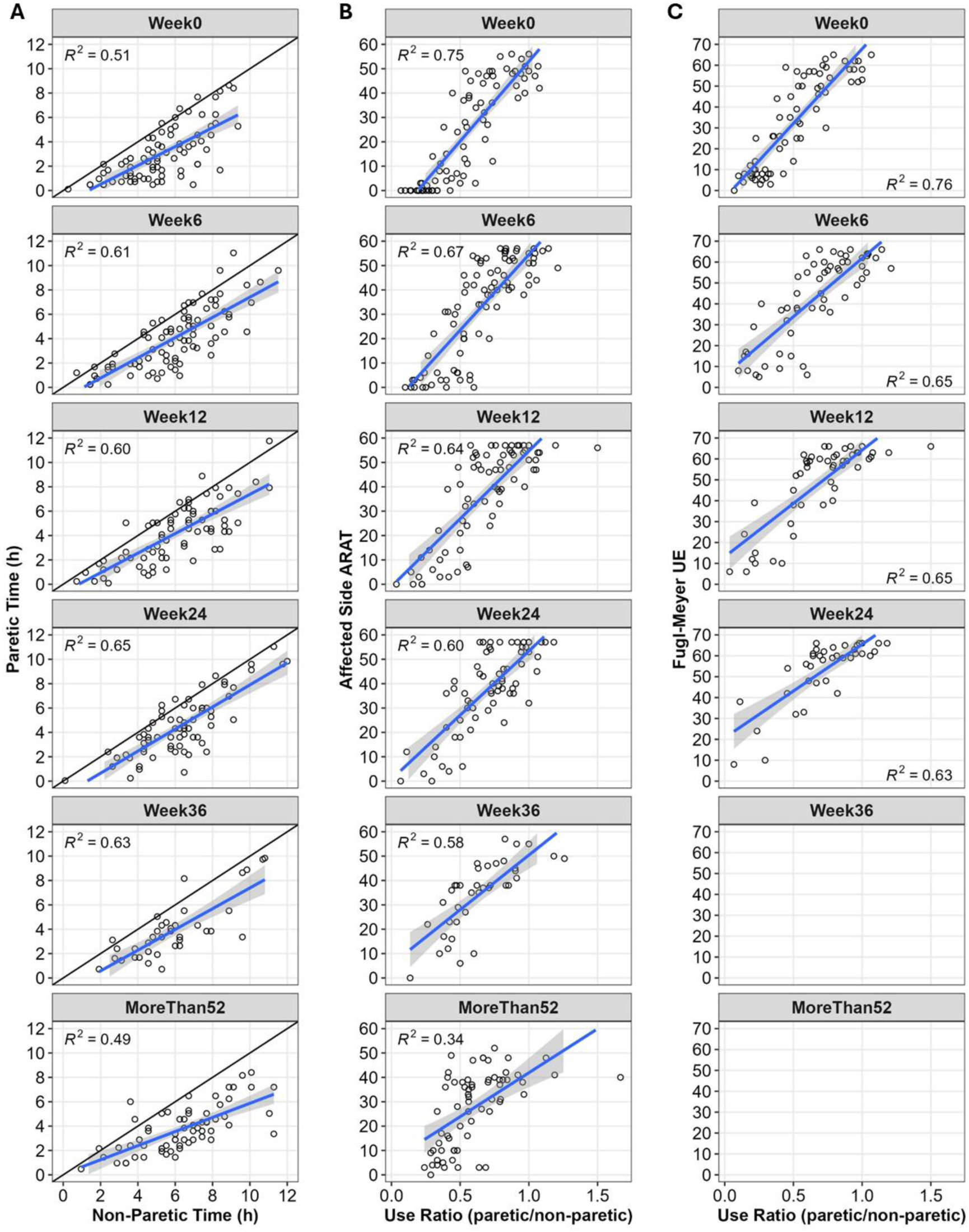
Associations between the paretic and non-paretic arm (**A**), ARAT and the use ratio (**B**), and the FM UE and the use ratio (**C**) at each of the different time points. Explained variance (r^2^) is shown for each comparison. Blue lines show the ordinary least squares regression fit, and the shaded region shows the 95% confidence interval. An additional black line of identity shows the hypothetical one-to-one relationship between the paretic and non-paretic arms. Plots show the first available observation for each person in each window of time, e.g., “Week0” = [0, 6) weeks post-stroke; “Week6” = [6,12) weeks post-stroke.

Comparing the use ratio to the ARAT (Figure 3B), we saw decreasing correlations between these variables over time: at [0, 6) weeks r = 0.87 and 95% CI=[0.80, 0.91]; at [6, 12) weeks r = 0.82 and 95% CI=[0.74, 0.88]; at [12, 24) weeks r = 0.80 and 95% CI=[0.70, 0.87]; at [24, 36) weeks r=0.77 and 95% CI=[0.66, 0.86]; at [36, 52) weeks r = 0.76 and 95% CI=[0.58, 0.86]; and at ≥52 weeks r =0.58 and 95% CI=[0.39, 0.72]. Comparing the use ratio to the FM UE (Figure 3C), we also saw decreasing correlations between these variables over time: from [0, 6) weeks r = 0.87 and 95% CI=[0.79, 0.92]; [6, 12) weeks r = 0.81 and 95% CI=[0.69, 0.88]; [12, 24) weeks r = 0.80 and 95% CI=[0.68, 0.88]; and to [24, 36) weeks r=0.79, and 95% CI=[0.63, 0.89]. Insufficient data were available after 36 weeks post-stroke for the FM-UE.

For both the FM-UE and the ARAT, we consistently saw very strong initial correlations with the use ratio that decreased over time. Importantly, these correlations were also much stronger for the use ratio than for the activity of the paretic hand or activity of the non-paretic hand alone (see Supplemental Figure ii and iii). Thus, it appears to be specifically the *asymmetry* in the activity of the upper-extremities that has a strong correspondence to in-clinic assessments of impairment (FM-UE) and the capacity for activity (ARAT), especially in the early weeks following stroke.

### 2. Associations in the Longitudinal Trajectories of Clinical Variables and the Use Ratio

Descriptive statistics for the longitudinal subset are provided in Table 2. The longitudinal subsample was demographically similar to the cross-sectional subsample, other than the difference in time poststroke, as by design the longitudinal sample included only participants enrolled <12 weeks post stroke: median = 1.3, IQR=[0.7, 4.2] weeks post-stroke.

**Table 2.** Descriptive statistics for the longitudinal cohort (N=98).

| Variable | Summary | Missing |
| --- | --- | --- |
| **Demographics** |  |  |
| Age at Enrollment | 64.5 [58.0, 71.0] | 0% (0) |
| Birth Sex |  | 0% (0) |
| Male | 59% (58) |  |
| Female | 41% (40) |  |
| Race |  | 0% (0) |
| White | 60% (59) |  |
| Black (or African American) | 39% (38) |  |
| Asian | 1% (1) |  |
| American Indian or Alaska Native | 0% (0) |  |
| Ethnicity |  | 0% (0) |
| Non-Hispanic/Non-Latine | 100% (98) |  |
| Hispanic/Latine | 0% (0) |  |
| Years of Education |  | 3% (3) |
| <High School | 30% (29) |  |
| Vocational/Associate's Degree | 9% (9) |  |
| Some College (no degree) | 32% (31) |  |
| Bachelor's Degree | 12% (12) |  |
| Masters/Doctoral/Professional Degree | 14% (14) |  |
| **Characterization of Stroke** |  |  |
| Stroke Type |  | 0% (0) |
| Ischemic | 83% (81) |  |
| Hemorrhagic | 16% (16) |  |
| Unknown | 1% (1) |  |
| Stroke Location |  | 13% (13) |
| Cortical | 51% (50) |  |
| Subcortical | 31% (31) |  |
| Cortical and Subcortical | 1% (1) |  |
| Posterior circulation/Cerebellar | 2% (2) |  |
| Other | 1% (1) |  |
| Number of Strokes |  | 1% (1) |
| First | 94% (92) |  |
| Second | 2% (2) |  |
| Third | 3% (3) |  |
| Affected Side |  | 0% (0) |
| Left | 39% (38) |  |
| Right | 61% (60) |  |
| Therapy/Rehab Location Type |  | 6% (6) |
| Inpatient Rehabilitation | 58% (57) |  |
| Outpatient Rehabilitation | 33% (32) |  |
| Skilled Nursing Facility | 1% (1) |  |
| Home Health | 2% (2) |  |
| **Prognostic Variables at Enrollment** |  |  |
| Time since Stroke (weeks) | 1.3 [0.7, 4.2] | 0% (0) |
| MoCA Total (/30) | 20.5 [16.0, 24.0] | 8% (8) |
| CES-Depression Total (/60) | 15 [6, 21] | 37% (36) |
| **Outcomes at Enrollment** |  |  |
| ARAT for most affected arm (/57) | 26.5 [3, 44.8] | 0% (0) |
| Fugl-Meyer Upper Extremity (/66) | 34 [9.5, 54.3] | 31% (30) |
| Non-paretic arm movement time (h) | 5.3 [3.9, 6.9] | 0% (0) |
| Paretic arm (h) | 2.6 [1.4, 4.6] | 0% (0) |
| Use Ratio (paretic/non-paretic) | 0.5 [0.3, 0.7] | 0% (0) |
| # of Observations per Person | 8 [5, 8] | *na* |

Note, summaries show either median [interquartile range] for continuous variables or % (n) for categorical variables. MoCA = Montreal Cognitive Assessment<sup>49</sup>, 18 to 25 is often considered “mild” impairment and 10 to 17 is considered “moderate impairment”; CES-Depression = Center for Epidemiological Studies Depression Scale<sup>50</sup>, scores >16 provide some evidence of depression with higher scores indicating more/greater severity of symptoms.

The estimated individual trajectories for each person are shown in Figure 4 for each of the three outcomes. The distributions and covariances for the different parameters are shown in Figure 5. Intercepts and slopes for weeks [0 to 5) are shown in Figure 5A and 5B, respectively. Slopes for weeks [5 to 11) are shown in Figure 5C. And slopes for after Week 11 are shown in Figure 5D. The diagonal panels in each figure show the univariate distributions for slopes and intercepts. The distribution of slopes (slope = change in points/week) tended to be most positive early, decline in Week 5, and generally flatten out after Week 11 for all measures. For the ARAT in Week 0, the mean change was 3.2 points/week; in Week 5 was 1.3 points/weeks; and in Week 11 was 0.13 points/week. Similarly, for the FM-UE at Week 0, the mean change was 2.7 points/week; in Week 5 was 1.3 points/week; and in Week 11 was 0.15 points/week. For the Use Ratio in Week 0, the mean change was 0.045 points/week; in Week 5 was 0.007 points/week; and in Week 11 was 0.002 points/week.

**Figure 4.**
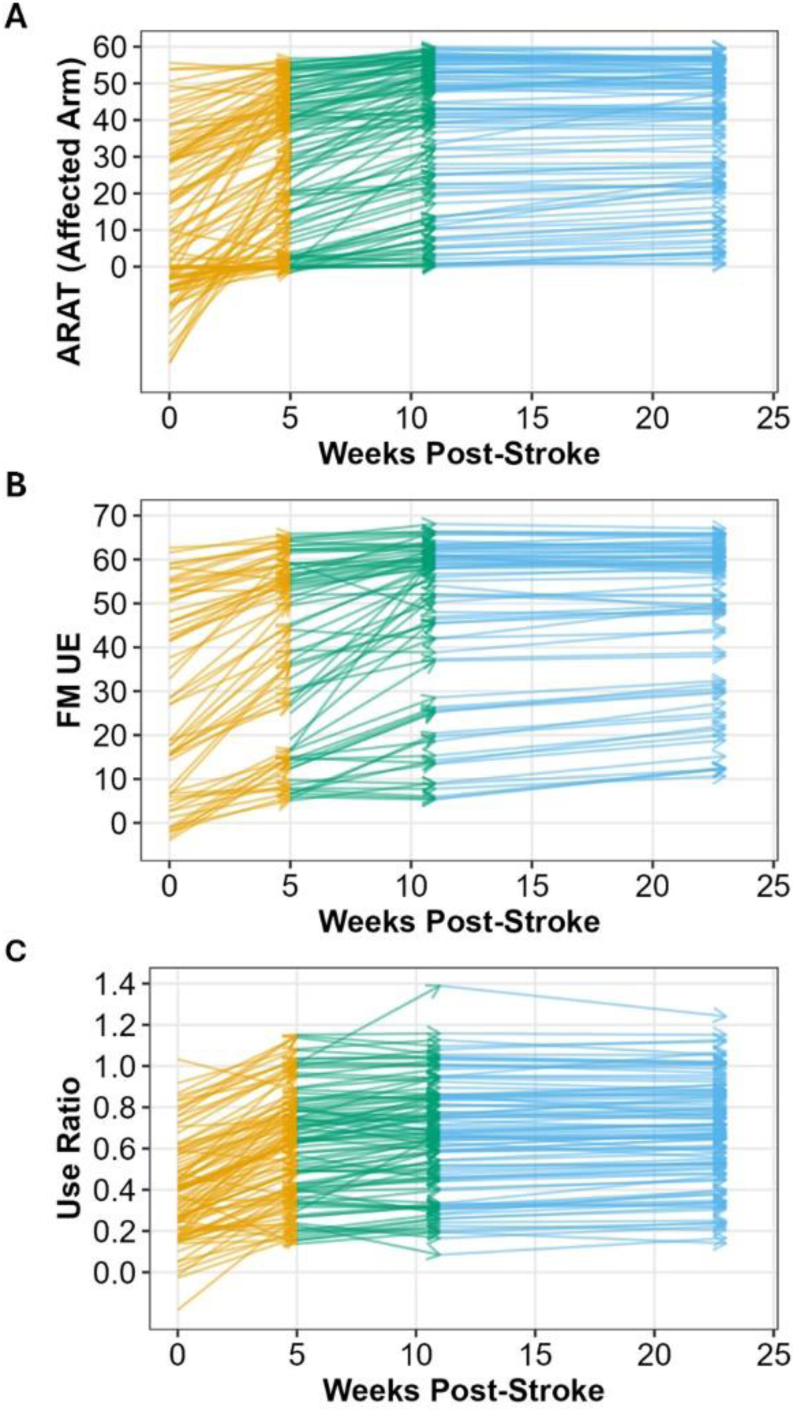
Estimated trajectories for the 2-knot spline model for each subject shown for the ARAT (A), the FM-UE (B), and the use ratio (C). Vectors are color-coded to highlight the different slopes at each point: orange = [0, 5) weeks; green = [5, 11) weeks; and blue = [11, 24) weeks.

**Figure 5.**
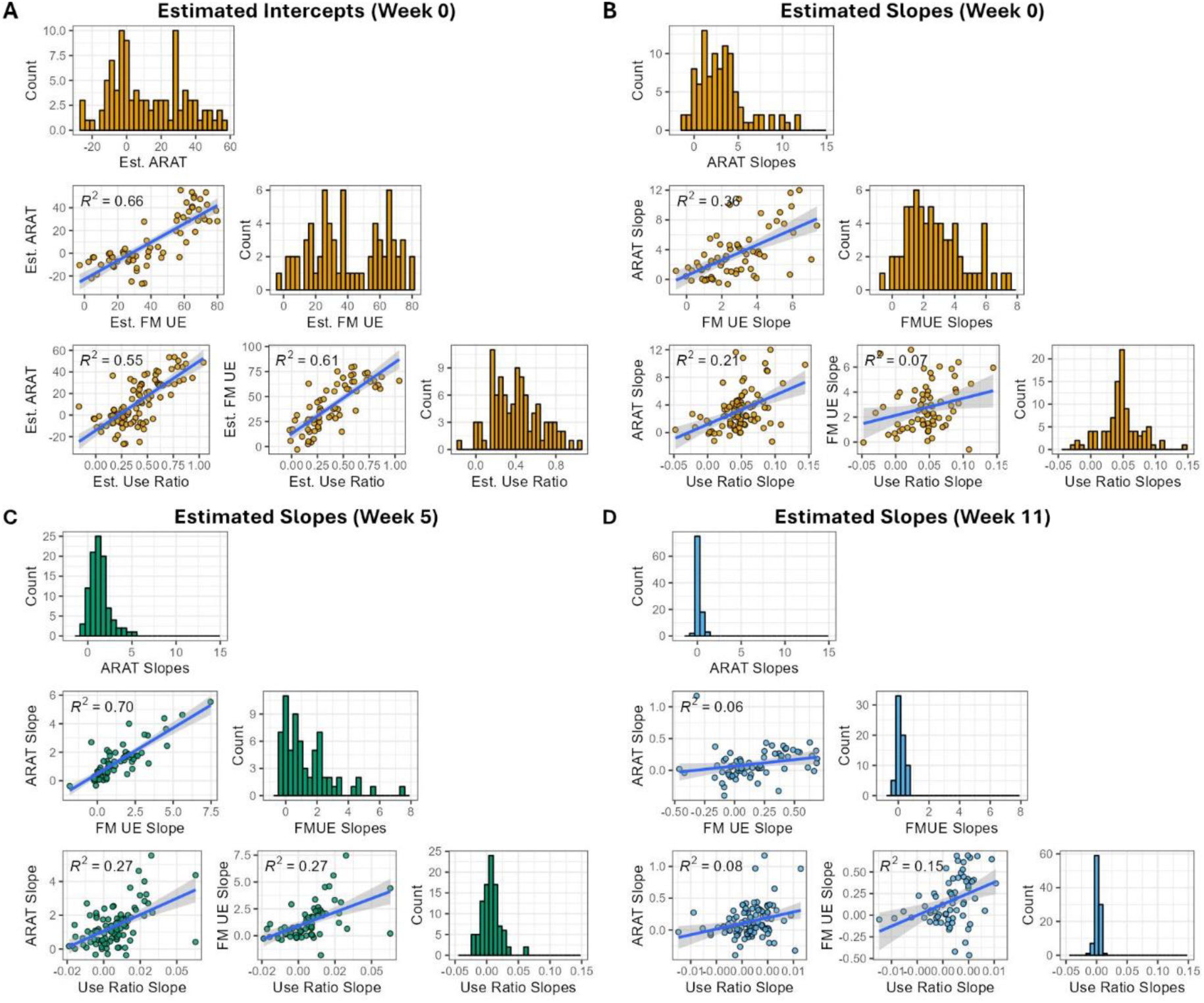
Associations between the components of individual trajectories across our primary outcomes. Intercepts and slopes from the 2-knot spline model are shown at Week 0 (A, B), slopes at Week 5 (C), and slopes are Week 11 (D).

Covariances between the trajectories for different outcome measures are shown in the off diagonals of each panel of Figure 5. These correlations were strongest for the intercepts (Figure 5A). The slopes for the different outcomes are positively associated in Weeks 0 to 5 (Figure 5B), and in Weeks 5 to 11 (Figure 5C). After Week 11, the correlations between trajectories were still positive, but greatly attenuated (Figure 5D). R-values and 95% confidence intervals for all coefficients at the different time points are shown in Table 3. These covariances show a similar pattern to the cross-sectional analyses (i.e., correlations between the measures decrease over time), but in more detail. Specifically, the longitudinal data allow us to break these trajectories into between-subject and within-subject agreement between variables. The positively correlated intercepts speak to *between-subject agreement*: if Person A has a low rank on the ARAT they also tend to have a low rank on the Use Ratio relative to their peers. In contrast, the positively correlated slopes speak to *within-subject agreement*: if the ARAT changes for Person A, then there is also a proportional change in the Use Ratio.

**Table 3.**
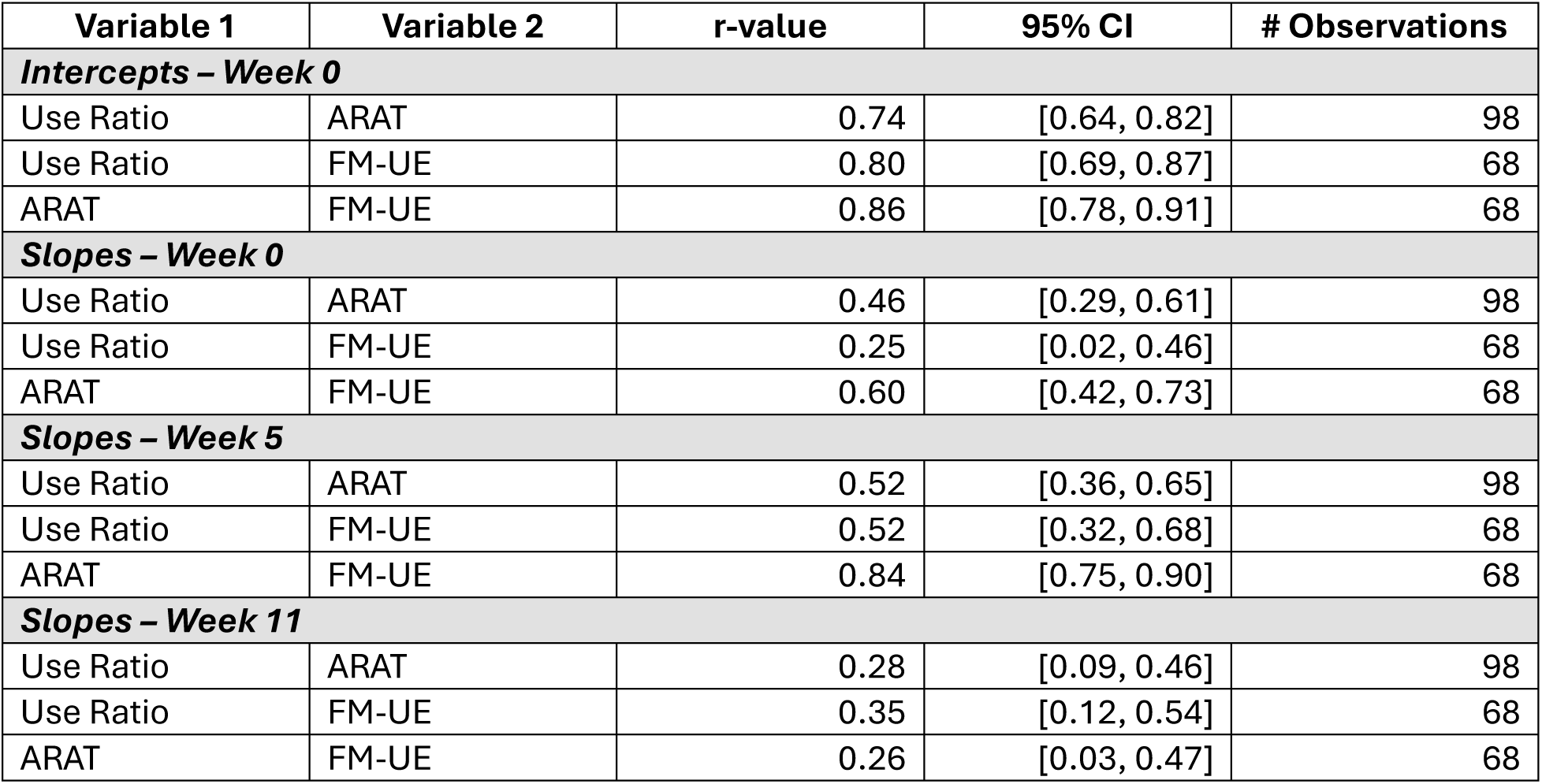
Correlations between components of the trajectory for each outcome at each time point.

| Variable 1 | Variable 2 | r-value | 95% CI | # Observations |
| --- | --- | --- | --- | --- |
| ***Intercepts – Week 0*** |  |  |  |  |
| Use Ratio | ARAT | 0.74 | [0.64, 0.82] | 98 |
| Use Ratio | FM-UE | 0.80 | [0.69, 0.87] | 68 |
| ARAT | FM-UE | 0.86 | [0.78, 0.91] | 68 |
| ***Slopes – Week 0*** |  |  |  |  |
| Use Ratio | ARAT | 0.46 | [0.29, 0.61] | 98 |
| Use Ratio | FM-UE | 0.25 | [0.02, 0.46] | 68 |
| ARAT | FM-UE | 0.60 | [0.42, 0.73] | 68 |
| ***Slopes – Week 5*** |  |  |  |  |
| Use Ratio | ARAT | 0.52 | [0.36, 0.65] | 98 |
| Use Ratio | FM-UE | 0.52 | [0.32, 0.68] | 68 |
| ARAT | FM-UE | 0.84 | [0.75, 0.90] | 68 |
| ***Slopes – Week 11*** |  |  |  |  |
| Use Ratio | ARAT | 0.28 | [0.09, 0.46] | 98 |
| Use Ratio | FM-UE | 0.35 | [0.12, 0.54] | 68 |
| ARAT | FM-UE | 0.26 | [0.03, 0.47] | 68 |

## Discussion

By leveraging a large, harmonized dataset of individuals with stroke who varied in initial severity and recovery trajectories, this study sheds new light on the utility of using scalable technologies to measure motor recovery post stroke. Broadly speaking the use ratio, measured via accelerometry in the home, demonstrated good concurrent validity with the ARAT and the FM-UE, measured in the clinic. In our cross-sectional analyses, hours of activity for the paretic arm were lower than the hours of activity for the non-paretic arm, as would be expected both from past research and the nature of hemiparesis following unilateral stroke.^51,52^ Critically, the correlation between hours of paretic and non-paretic limb activity were also relatively stable, suggesting that the use ratio is consistently capturing individual differences in limb asymmetry over time.^19^ In both the cross-sectional and the longitudinal analyses, the use ratio was positively correlated with the ARAT and the FM-UE. Cross-sectionally, between people, *a person* with a higher use ratio reliably had a higher ARAT and FM-UE. Longitudinally, within a person, *a change* in the use ratio was also associated with corresponding changes in the ARAT and the FM-UE. Critically however, we show for the first time that the strength of these correlations decreased over time (albeit remaining positive) for both the cross-sectional and longitudinal analyses.

The attenuation of these correlations over time may be explained by what we know about the different domains of the ICF and the time-course of recovery following a stroke.^19^ Especially in the early phases following a stroke (e.g., <11 weeks), biological recovery and thus body functions may be the rate limiting factor on what a person can do.^47,53^ As recovery progresses however, behavioral *compensation* may enable a person to perform activities despite impairments in body function.^54^ Similarly, environmental and personal factors will start to play a larger role in a person’s performance of different activities in daily life and the person’s ability to participate in different social roles (e.g., cognitive and affective impairments have a larger moderating effect on motor behavior when a person returns home^55^).

The concurrent validity of the use ratio with the ARAT and the FM-UE strongly suggests that scalable and efficient actigraphy is a solution for capturing phenotypes in motor recovery, but the timing of the assessment matters. First, within the early phase of recovery we saw three relevant timepoints that need to be assessed: a baseline within the first week following the stroke, an inflection-point around 5 weeks after the stroke, and an inflection-point around 11 weeks after the stroke. These timepoints captured individual variation in trajectories for three different outcome measures. The timepoints also align well with known “milestones” in the neurobiology of recovery.^45,46^ Taken together, this suggests that at least three assessments around 0, 5, and 11 weeks will be important for establishing informative motor recovery phenotypes. Second, we need to consider the divergence between the use ratio, ARAT, and FM-UE later in recovery. Beyond 11 weeks, actigraphy appears to provide related, but more distinct information, perhaps reflecting the influence of more “person-level” factors (e.g., psychological, sociological, and environmental factors^19^). Although this means that the use ratio may become less representative of body functions and the biology of recovery, it also means that actigraphy may be more reflective of participation in social roles (e.g., family, work) which are often rated as very important by patients.^56,57^

Changes in the correlation with the ARAT and FM-UE over time reflects strengths and limitations of actigraphy for stroke recovery phenotyping. As a strength, motor deficits are: 1) highly prevalent in people with stroke^58,59^; 2) generally have a shorter recovery time scale than other domains^60^; 3) of substantial concern to people living with stroke^61,62^; and 4) are one of the easier domains of recovery to measure.^63^ The use ratio provides a single outcome that can be consistently tracked across different phases of motor recovery in large samples of people with stroke, using scalable technology that minimizes burden on both participants and research staff. As a limitation, especially at later stages of recovery, researchers may need to collect additional information about personal, social, and environmental factors to contextualize actigraphy (e.g., major depression affects how much a person moves, even if they have the capacity for greater levels of activity). As technology and research progress, an ideal vision is to remotely collect a battery of outcomes – motoric, linguistic, cognitive, affective – and their moderators, across many domains of recovery.

## Conclusion

This is the largest study to date on the measurement properties of actigraphy for capturing motor recovery following a stroke. The use ratio, calculated from bilateral wrist worn sensors, has very good concurrent validity with in-clinic measures of impairment (FM-UE) and activity capacity (ARAT) in a large heterogenous sample of human adults with stroke. Positive associations between these measures were found both cross-sectionally (i.e., between person *differences*) and longitudinally (i.e., within person *changes*), but the strength of these associations changed over time. Although generally positive in all phases of recovery, these associations were strongest in the first few weeks after a stroke, but the use ratio generally started to diverge from other measures around 11-weeks post-stroke. We speculate that changes in the strength of these associations reflect changes in the underlying factors that shape spontaneous motor behavior. Early in the recovery process, biological recovery is likely the rate limiting factor on activity, creating greater convergence between measures. Later in the recovery process, personal, social, and environmental factors likely have a greater influence on upper extremity activities, creating more divergence between these measures which tap into related but distinct constructs: FM-UE (body function), ARAT (activity capacity in the clinic), and the use ratio (activity performance in daily life). In summary, the use ratio is a scalable, efficient, clinically valid outcome for measuring upper extremity motor recovery following stroke.

## Data Availability

All data for this study are publicly available from the NICD Data and Specimen Hub. All code the for the analyses are openly available from the lead author's github repository. All relevant URLs are provided in the text.

https://doi.org/10.57982/fayx-p832

https://doi.org/10.57982/72z7-m179

