## Supplemental Materials for "Validation of real-world actigraphy to capture post-stroke motor recovery"

### Supplemental Appendix

#### Supplemental Figure i.

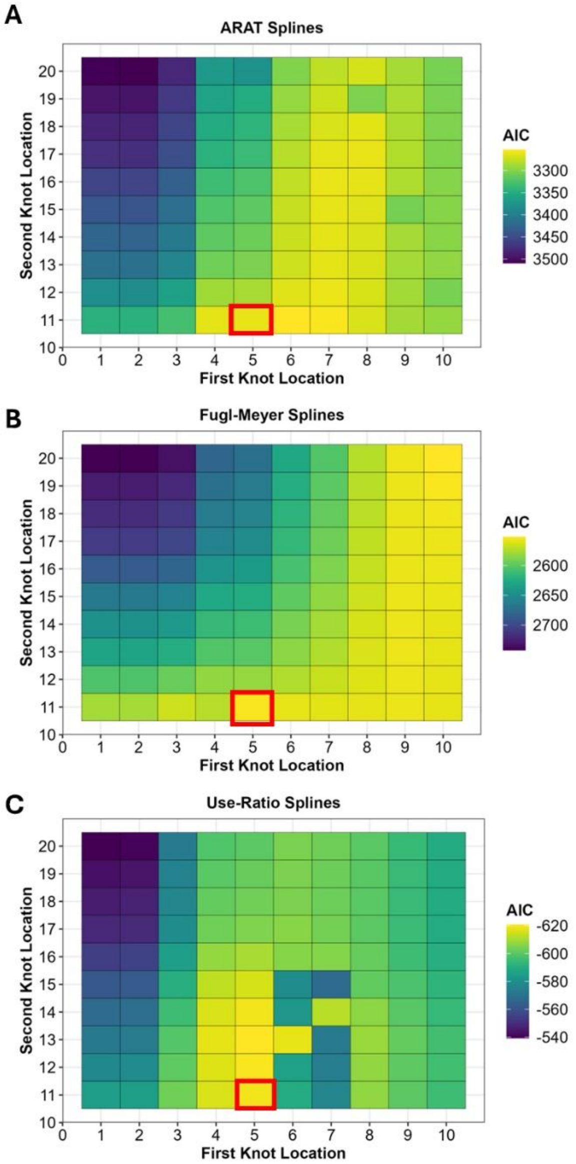

**Supplemental Figure ii.**

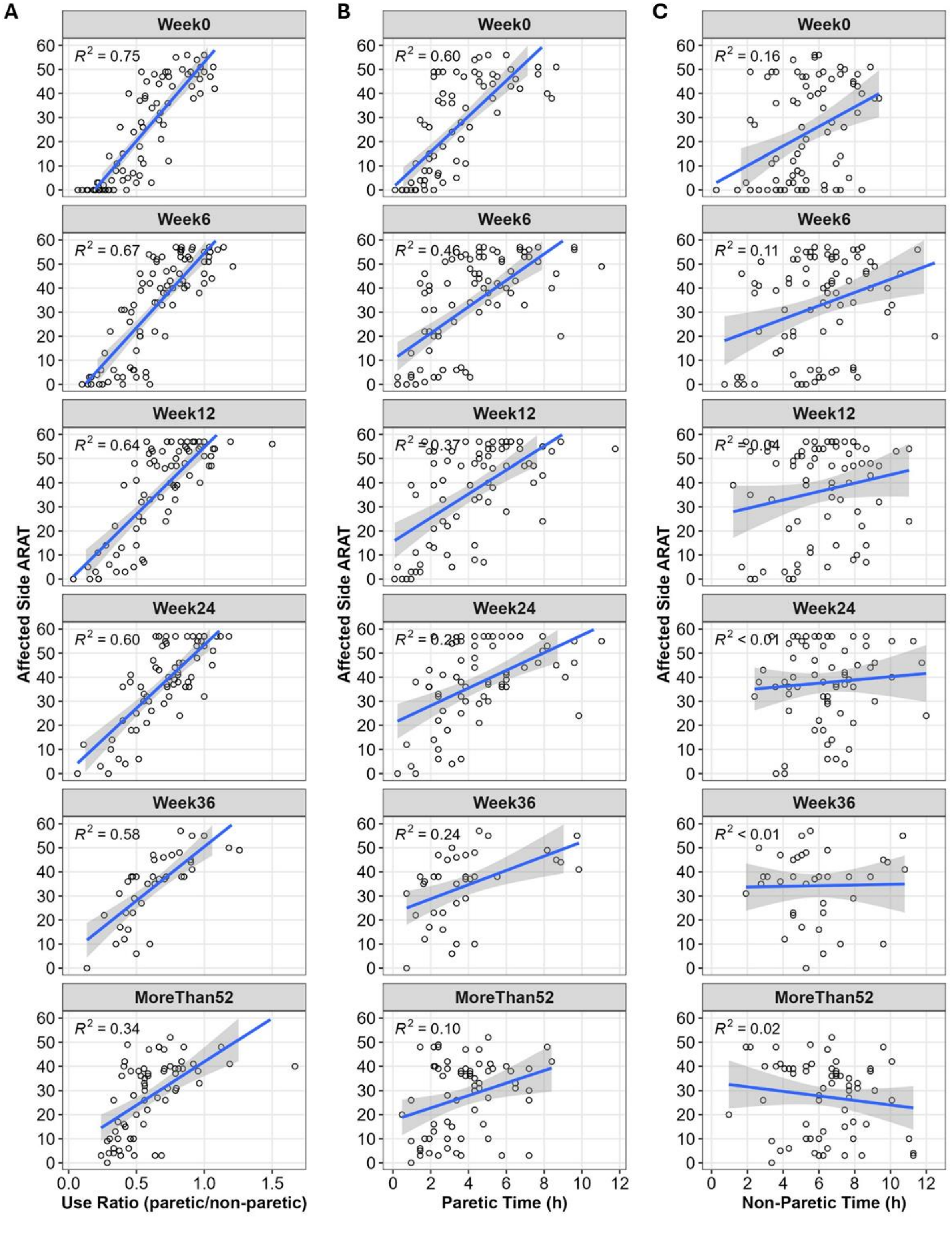

**Supplemental Figure iii.**

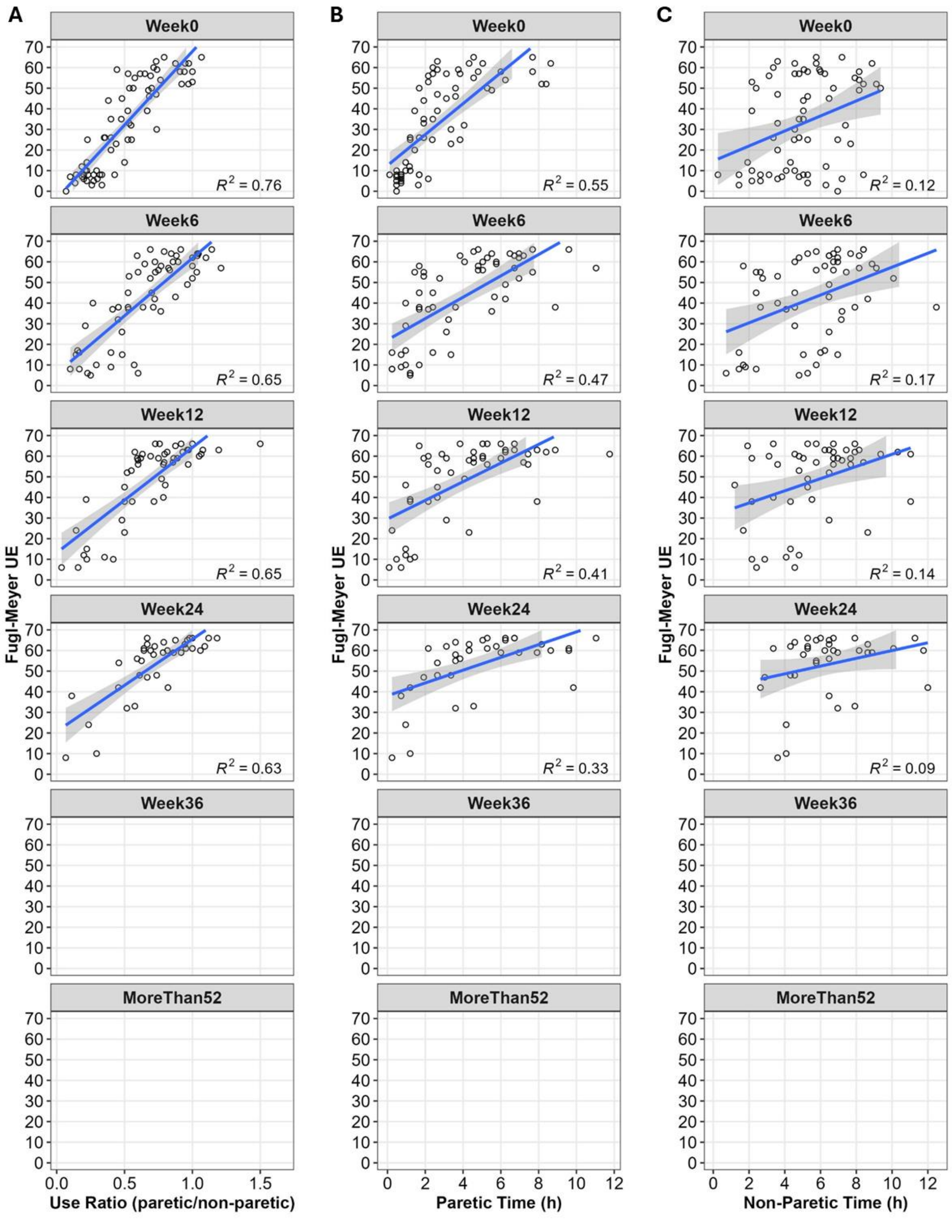

**Supplemental Table i.** Model fit comparisons for the ARAT

| Model | n par | AIC | BIC | logLik | deviance | $\chi^2$ Decrease | df | P($\chi^2$) |
|---|---|---|---|---|---|---|---|---|
| Intercept Only | 3 | 3777.1 | 3789.8 | -1885.6 | 3771.1 | | | |
| Linear Time | 6 | 3537.5 | 3562.7 | -1762.8 | 3525.5 | 245.634 | 3 | <0.001 |
| Inverse Square | 6 | 3379.9 | 3405.1 | -1684.0 | 3367.9 | 157.594 | 0 | *na* |
| Inverse Cubic | 6 | 3479.8 | 3505.1 | -1733.9 | 3467.8 | (increased) | 0 | *na* |
| 3-Param. Exponential | 9 | 3283.5 | 3325.5 | -1631.8 | 3263.5 | 204.3 | 3 | <0.001 |
| Quadratic Time | 10 | 3373.1 | 3415.1 | -1676.5 | 3353.1 | (increased) | 1 | *na* |
| Cubic Time | 11 | 3338.1 | 3384.3 | -1658.0 | 3316.1 | 37.016 | 1 | <0.001 |
| ***2-Knot Spline†*** | ***15*** | ***3261.9*** | ***3325.0*** | ***-1615.9*** | ***3231.9*** | ***84.198*** | ***4*** | ***<0.001*** |

Note, all models included random slopes for all time parameters. The 2-knot spline shown refers to overall best fitting spline with knots at 5- and 11-weeks. AIC = Akaike's information criterion; BIC = Bayesian Information Criterion; loglik = log(Likelihood); df = degrees of freedom; p($\chi^2$) = p-value from Wald's test for the change in deviance compared to previous model. † = best fitting model based on AIC.

**Supplemental Table ii.** Model fit comparisons for the FM-UE

| Model | n par | AIC | BIC | logLik | deviance | $\chi^2$ Decrease | df | P($\chi^2$) |
|---|---|---|---|---|---|---|---|---|
| Intercept Only | 3 | 2969.9 | 2981.8 | -1481.9 | 2963.9 | | | |
| Linear Time | 6 | 2772.2 | 2796 | -1380.1 | 2760.2 | 203.696 | 3 | < 2.2e-16 |
| Inverse Square | 6 | 2675.1 | 2698.9 | -1331.5 | 2663.1 | 97.12 | 0 | *na* |
| Inverse Cubic | 6 | 2759.2 | 2783 | -1373.6 | 2747.2 | (increased) | 0 | *na* |
| 3-Param. Exponential | 6* | 2578.9 | 2606.8 | -1282.5 | 2564.9 | 182.2 | 0 | *na* |
| Quadratic Time | 10 | 2619.8 | 2659.5 | -1299.9 | 2599.8 | (increased) | 4 | *na* |
| Cubic Time | 11 | 2602.3 | 2646 | -1290.1 | 2580.3 | 19.535 | 1 | 9.88E-06 |
| ***2-Knot Spline†*** | ***15*** | ***2554.1*** | ***2613.7*** | ***-1262.1*** | ***2524.1*** | ***56.15*** | ***4*** | ***1.87E-11*** |

Note, all models included random slopes for all time parameters. The 2-knot spline shown refers to overall best fitting spline with knots at 5- and 11-weeks. AIC = Akaike's information criterion; BIC = Bayesian Information Criterion; loglik = log(Likelihood); df = degrees of freedom; p($\chi^2$) = p-value from Wald's test for the change in deviance compared to previous model. * = model failed to converge with random-effect of the rate parameter, so was refit with only a fixed rate parameter. † = best fitting model based on AIC.

**Supplemental Table iii.** Model fit comparisons for the Use Ratio

| Model | n par | AIC | BIC | logLik | deviance | $\chi^2$ Decrease | df | $P(\chi^2)$ |
|---|---|---|---|---|---|---|---|---|
| Intercept Only | 3 | -459.45 | -446.82 | 232.73 | -465.45 | | | |
| 3-Param. Exponential | 4* | -548.3 | -527.2 | 279.10 | -558.28 | 92.8 | 1 | < 2.2e-16 |
| Linear Time | 6 | -533.04 | -507.76 | 272.52 | -545.04 | (increased) | 2 | *na* |
| Inverse Square | 6 | -608.13 | -582.85 | 310.06 | -620.13 | 75.0868 | 0 | |
| Inverse Cubic | 6 | -587.68 | -562.4 | 299.84 | -599.68 | (increased) | 0 | |
| Quadratic Time | 10 | -588.96 | -546.83 | 304.48 | -608.96 | 9.2819 | 4 | 0.054427 |
| Cubic Time | 11 | -597.73 | -551.39 | 309.87 | -619.73 | 10.7704 | 1 | 1.03E-03 |
| ***2-Knot Spline†*** | ***15*** | ***-619.19*** | ***-556*** | ***324.60*** | ***-649.19*** | ***29.4616*** | ***4*** | ***6.30E-06*** |

Note, all models included random slopes for all time parameters. The 2-knot spline shown refers to overall best fitting spline with knots at 5- and 11-weeks. AIC = Akaike's information criterion; BIC = Bayesian Information Criterion; loglik = log(Likelihood); df = degrees of freedom; $p(\chi^2)$ = p-value from Wald's test for the change in deviance compared to the previous model. * = model failed to converge with random-effects of the rate and change parameters, so was refit with only a random effect on the asymptote. † = best fitting model based on AIC.

**Supplemental Table iv.** Details of the best fitting spline model for the ARAT.

```
     AIC      BIC   logLik deviance df.resid
  3261.9   3325.0  -1615.9   3231.9      481

Scaled residuals:
    Min      1Q  Median      3Q     Max
-3.4678 -0.3429  0.0152  0.2958  3.0871

Random effects:
 Groups    Name            Variance Std.Dev. Corr
 SubIDName (Intercept)     553.605  23.529
           WeeksPostStroke  13.226   3.637   -0.56
           knot05           11.695   3.420    0.43 -0.93
           knot11            1.645   1.283    0.43 -0.47  0.13
 Residual                    7.419   2.724
Number of obs: 496, groups:  SubIDName, 98

Fixed effects:
                Estimate Std. Error      df t value Pr(>|t|)
(Intercept)      12.7206     2.8857 85.6438   4.408 3.01e-05 ***
WeeksPostStroke   3.1852     0.5159 68.0570   6.174 4.19e-08 ***
knot05           -1.8513     0.5306 67.2708  -3.489  0.00086 ***
knot11           -1.2056     0.1836 81.1598  -6.566 4.54e-09 ***
---
Signif. codes:  0 ‘***’ 0.001 ‘**’ 0.01 ‘*’ 0.05 ‘.’ 0.1 ‘ ’ 1
```

**Supplemental Table v.** Details of the best fitting spline model for the FM-UE.

```
     AIC      BIC   logLik deviance df.resid 
  2554.1   2613.7  -1262.1   2524.1      378 

Scaled residuals: 
    Min      1Q  Median      3Q     Max 
-3.1768 -0.3573  0.0209  0.3821  3.3408 

Random effects:
 Groups    Name            Variance Std.Dev. Corr             
 SubIDName (Intercept)     622.746  24.955                    
           WeeksPostStroke   5.707   2.389   -0.54            
           knot05            6.314   2.513    0.22 -0.73      
           knot11            3.833   1.958    0.30 -0.25 -0.47
 Residual                    7.847   2.801                    
Number of obs: 393, groups:  SubIDName, 68

Fixed effects:
                Estimate Std. Error      df t value Pr(>|t|)    
(Intercept)      24.7254     3.2574 66.1576   7.591 1.41e-10 ***
WeeksPostStroke   2.7116     0.4118 57.3172   6.585 1.52e-08 ***
knot05           -1.4258     0.4737 52.5736  -3.010 0.004008 ** 
knot11           -1.1319     0.2939 57.4449  -3.851 0.000298 ***
---
Signif. codes:  0 ‘***’ 0.001 ‘**’ 0.01 ‘*’ 0.05 ‘.’ 0.1 ‘ ’ 1
```

**Supplemental Table vi.** Details of the best fitting spline model for the Use Ratio.

```
     AIC      BIC   logLik deviance df.resid 
  -619.2   -556.0    324.6   -649.2      484 

Scaled residuals: 
    Min      1Q  Median      3Q     Max 
-3.6433 -0.4583 -0.0067  0.3897  4.0693 

Random effects:
 Groups    Name            Variance  Std.Dev. Corr             
 SubIDName (Intercept)     0.0808610 0.28436                   
           WeeksPostStroke 0.0019462 0.04412  -0.43            
           knot05          0.0024237 0.04923   0.31 -0.90      
           knot11          0.0006486 0.02547  -0.02  0.18 -0.59
 Residual                  0.0054398 0.07375                   
Number of obs: 499, groups:  SubIDName, 98

Fixed effects:
                 Estimate Std. Error        df t value Pr(>|t|)    
(Intercept)      0.395352   0.042263 74.221063   9.355 3.47e-14 ***
WeeksPostStroke  0.045277   0.008431 68.480103   5.370 1.01e-06 ***
knot05          -0.038468   0.009880 72.821508  -3.893 0.000217 ***
knot11          -0.005270   0.004160 84.278495  -1.267 0.208719    
---
Signif. codes:  0 ‘***’ 0.001 ‘**’ 0.01 ‘*’ 0.05 ‘.’ 0.1 ‘ ’ 1
```
